# Strong early impact of letrozole on ovulation induction outperforms clomiphene citrate in polycystic ovary syndrome: a systematic review with meta-analysis

**DOI:** 10.1101/2023.09.25.23296113

**Authors:** Rita Zsuzsanna Vajna, András Mihály Géczi, Fanni Adél Meznerics, Nándor Ács, Péter Hegyi, Emma Zoé Feig, Péter Fehérvári, Szilvia Kiss-Dala, Szabolcs Várbíró, Judit Réka Hetthessy, Levente Sára

**Author notes:** Corresponding author, (LS). Equal contribution, therefore, the authors should be considered as co-last authors (Judit Réka Hetthessy, Levente Sára).

## Abstract

**Background:** Polycystic ovary syndrome is one of the most frequent endocrinological problems causing infertility in women worldwide. The main problem in these women is hyperandrogenism and/or chronic oligo/anovulation, which leads to infertility. In this systematic review and meta-analysis, we aimed to investigate the efficacy of a relatively new drug for ovulation induction, letrozole, by comparing it to the first line of treatment for ovulation induction, clomiphene citrate.

**Methods:** A literary search was conducted in three databases and included randomized clinical trials comparing letrozole and clomiphene citrate for ovulation induction for women with polycystic ovary syndrome. The diagnosis of polycystic ovary syndrome was determined according to the Rotterdam criteria. We pooled data using a random-effects model.

**Results:** Our search provided a total of 1,994 articles, of which we included 25 studies. In the letrozole group, endometrial thickness was significantly higher (Mean Difference=1.70, Confidence Interval: 0.55-2.86; Heterogeinity: I^2^=97%, p-value=0.008); odds for ovulation (Odds Ratio=1.8, Confidence Interval: 1.21-2.69; Heterogeinity: I^2^=51%, p-value=0.010) and pregnancy (Odds Ratio=1.96, Confidence Interval: 1.37-2.81; Heterogeinity: I^2^=32%, p-value=0.002) were significantly higher; the resistance index of subendometrial arteries was significantly lower (Mean Difference=-0.15, Confidence Interval: -0.27- -0.04; Heterogeneity: I^2^=92%, p-value=0.030).

**Conclusion:** Women with polycystic ovary syndrome treated with letrozole for ovulation induction had higher ovulation and pregnancy rates, their endometrium became thicker, the resistance index of subendometrial arteries was lower. The lower resistance index of the subendometrial arteries can improve intrauterine circulation, which may provide better circumstances for embryo implantation and development.

## Introduction

Polycystic ovary syndrome (PCOS) is one of the most frequent endocrinological problems causing infertility in women worldwide [1–4]; the prevalence of this disease is between 9 and 18% [5]. The main features of the disease are hyperandrogenism and/or chronic oligo/anovulation, which leads to infertility [6, 7].

Clomiphene citrate (CC) has been used since 1960 as a first-line medication for ovulation induction (OI) for women with PCOS [8, 9]. However, it has some unpleasant and non-dose-dependent side effects, such as hot flashes, increase in ovarian size, bloating, nausea and vomiting, breast sensitivity and pain, headaches, hair loss, insomnia, and depression [10].

Because of the longer half-life of CC, pregnancy may not occur despite ovulation, perhaps due to its antiestrogenic effects on the endocervix and the endometrium [5, 8, 11].

Letrozole (LE) is a third-generation aromatase inhibitor, an approved adjuvant when treating estrogen receptor-positive breast cancer. [12–15]. LE does not show anti-estrogenic effects [16, 17] and has minor side effects such as leg cramps and headaches [18, 19]. LE is cleared from the circulation faster, with a shorter half-life compared to CC [8]. The aromatase inhibition develops, endometrium receptors are up-regulated and rapid endometrial growth is observed without adverse effects on endometrium receptivity [8, 15]. Better blood supply to the sub-endometrial halo and to the endometrium results in a thicker endometrial wall, thus maximizing the odds of pregnancy [20].

Despite its many beneficial properties, LE is not yet included in the first-line therapy of PCOS patients and little information is available on its effectiveness. Therefore, we aimed to investigate the efficacy of LE on ovulation rate, endometrial receptivity, and pregnancy rate in women with PCOS compared to the effects of CC by performing a systematic review and meta-analysis.

## Methods

We describe our systematic review and meta-analysis based on the recommendations of the PRISMA (Preferred Reporting Items for Systematic Reviews and Meta-Analyses) 2020 guidelines (**S2 Table**) [21], while we followed the Cochrane Handbook’s recommendations for Systematic Reviews of Interventions Version 6.1.0 [22]. The protocol of the study was registered on PROSPERO (registration number CRD42022376611; see https://www.crd.york.ac.uk/PROSPERO); the protocol was strictly adhered to, additionally, we conducted a post-hoc analysis including 12 additional outcomes indicated in the population-intervention-control-outcome (PICO) to provide a comprehensive overview of all potentially important outcomes.

### Literature search and eligibility criteria

We completed our systematic literature search in three medical databases: MEDLINE (via PubMed), Cochrane Library (CENTRAL), Embase, from inception to 21 November 2022.

We applied the query ‘(polycystic ovarian syndrome OR PCOS OR Stein-Leventhal syndrome) AND (letrozole OR aromatase inhibitor)’ to all fields in the search engines. Neither language nor other restrictions were imposed.

Randomized clinical trials (RCTs) comparing LE to CC were included for patients diagnosed with PCOS based on Rotterdam criteria (at least two years after menarche) [23], comparing LE to CC were included. The following PICO agenda was applied:

P: women with PCOS

**I: LE**

**C: CC**

O**: outcomes according to the protocol**: endometrial thickness (ET); number of dominant follicles; ovulation rate; pregnancy rate; endometrial volume; **additional outcomes:** endometrial pattern- and echogenicity; diameter of dominant follicles; rate of mono-and multifollicular development; single and multiple pregnancy rate; live birth rate; miscarriage rate; prevalence of ectopic pregnancies; number of fetal anomalies; endometrial vascularization index, flow index, vascularization flow index and detection rate of endometrial-subendometrial blood flow; resistance index (RI) and pulsatility index (PI) of subendometrial and uterine arteries; systolic velocity/diastolic velocity of subendometrial arteries; biomarker /vascular endothelial growth factor and integrin alpha vß3/ concentrations in uterine fluid

Reviews, cases series, cases reports; studies including patients without PCOS diagnosis, girls before menarche, and those within two years past menarche were excluded from the study.

### Study selection and data collection

The selection was performed by two independent reviews (RZSV and EZF). Data from the eligible articles were collected by two authors (RZSV and AMG) independently. The following data were extracted about the study: first author; study type; the year of publications; study population; study period. The following data were extracted about patients studied: age; body mass index; duration of infertility; if ovulation was supported with human chorionic gonadotropin or the luteal phase with progesterone; the time interval for endometrial testing in relation to the cycle; the dose of LE or CC and the time of their administration in relation to the cycle, and data on the outcomes.

### Study risk of bias assessment

Two authors (RZSV and AMG) performed the risk of bias assessment independently. Risk of bias was measured using the Risk of Bias Tool 2 [23], as recommended by the Cochrane Collaboration . The quality assessment of the studies included was accomplished according to the “Grades of Recommendation, Assessment, Development, and Evaluation (GRADE)” workgroup with GRADE-Pro, as recommended by the Cochrane Collaboration [24].

### Synthesis methods

Statistical analysis was conducted using the R 4.1.2. [25] and meta [26] and dmetar packages [27].

Letrozole and clomiphene citrate were compared by calculating the mean differences of endometrial thickness, number of dominant follicles, diameter of dominant follicles, RI of subendometrial artery and PI of subendometrial artery between the intervention and the control groups. Ovulation rate, pregnancy rate, single pregnancy rate, frequency of miscarriages, frequency of monofollicular and mutlifollicular development were also examined. The following outcomes were also analyzed separately in patients who ovulated only: ET, number of dominant follicles, and pregnancy rate.

For binary outcomes, odds ratios with 95% confidence intervals (CI) were used as outcome measures. The number of patients and events was extracted or calculated. The results are presented as the odds of an event in the letrozole group compared to the odds of the same event in the clomiphene citrate group.

For continuous data, differences in mean values were used with 95% confidence intervals.

To calculate mean differences, sample sizes, mean and standard deviation values were extracted from the studies. Mean differences were calculated by extracting the mean values of the clomiphene citrate group from the mean values of the letrozole group.

Pooled OR was calculated with the Mantel-Haenszel method [28] and Robins, Greenland, and Breslow [29]. The exact Mantel-Haenszel method [28] (without continuity correction) was used to handle zero cell counts as recommended by Cooper, Hedges, and Valentine [30] and J. Sweeting, J. Sutton, and C. Lambert [31]. Confidence intervals were created with the Paule-Mandel method [32], recommended by Veroniki et al. [33]. In case of 0 cell counts, individual study odds ratios with 95% confidence intervals were calculated by adding 0.5 as continuity correction (it was used only for visualization on forest plots).

Pooled mean differences were computed with the inverse variance method.

The restricted maximum-likelihood estimator was used with the Q profile method for confidence intervals by Harrer et al. [34] and Veroniki et al. [33]. Hartung-Knapp adjustments were also applied [35] and IntHout, Ioannidis, and Borm [36].

Between-study heterogeneity was assessed with the Higgins and Thompson’s I² [37] statistic and the Cochrane Q test recommended by Harrer et al. [34]. The I 2 statistic shows what percentage of heterogeneity cannot be explained by random chance. Heterogeneity is considered substantial if I 2 exceeds 75%.

### Publication bias and heterogeneity

Publication bias was measured using funnel plots and Egger’s test [38, 39]. In case of endometrial thickness and pregnancy rate. The funnel plots do not indicate publication bias and the result of Egger’s test was not significant, which means publication bias could not be detected.

In case of endometrial thickness, number of dominant follicles and diameter of dominant follicles, RI of subendometrial artery and PI of subendometrial artery substantial heterogeneity could be observed. Other outcome measures showed low or moderate heterogeneity.

## Results

### Search and selection

We found 1,994 articles as a result of our systematic search. After duplicate removal, title and abstract selection was performed on 1274 articles. Finally, we included 22 [1, 4, 15, 16, 19, 40–55] studies after full-text selection, and three additional studies based on citation search [5, 6, 56]. The details of the search and selection process are visualized in **Fig 1**.

**Fig 1.**
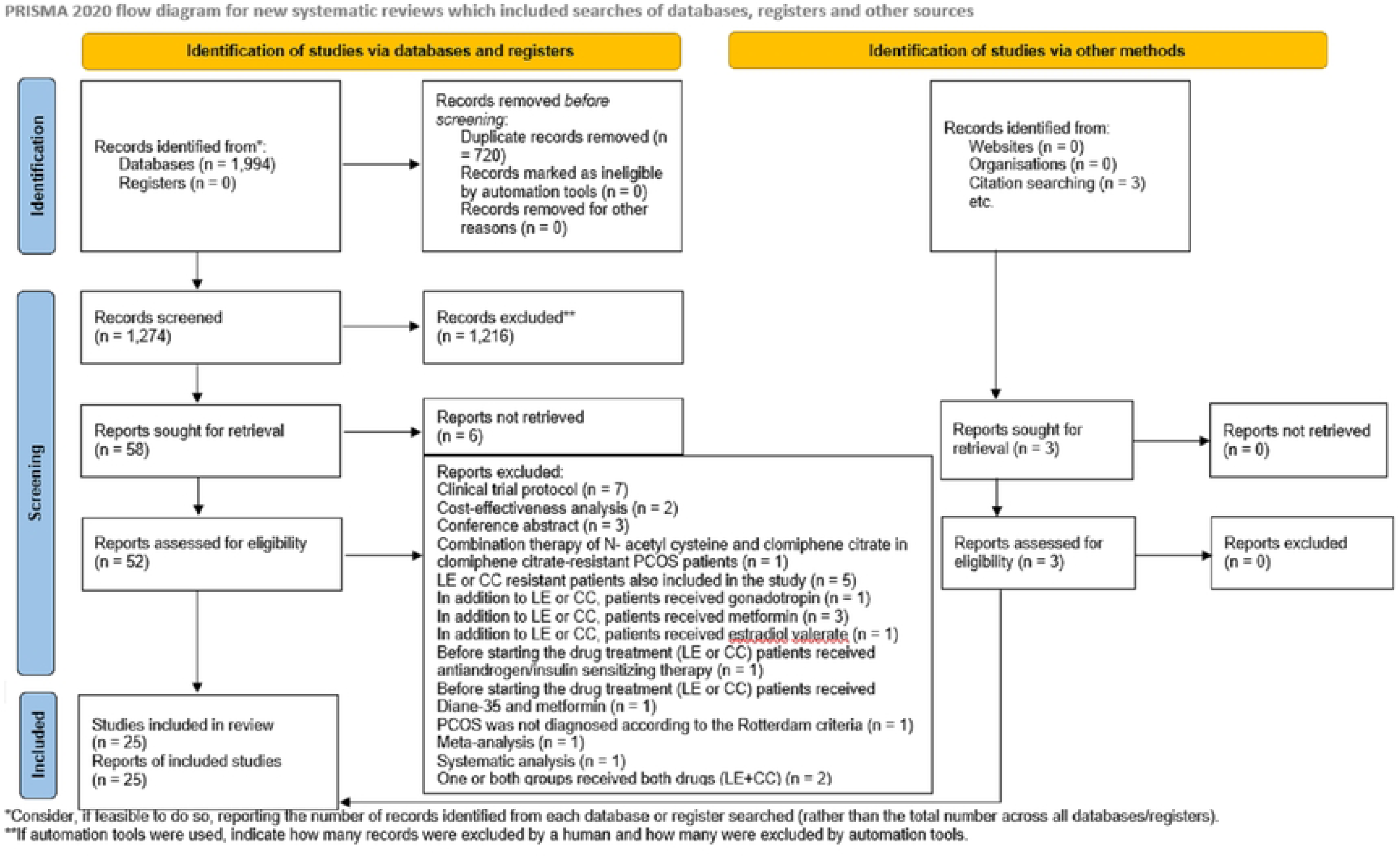
PRISMA (Page et al., 2021) flow diagram.

### Characteristics of the included studies

Baseline and patient characteristics of the studies included in the systematic review and meta-analysis are detailed in **Table 1**.

**Table 1.**
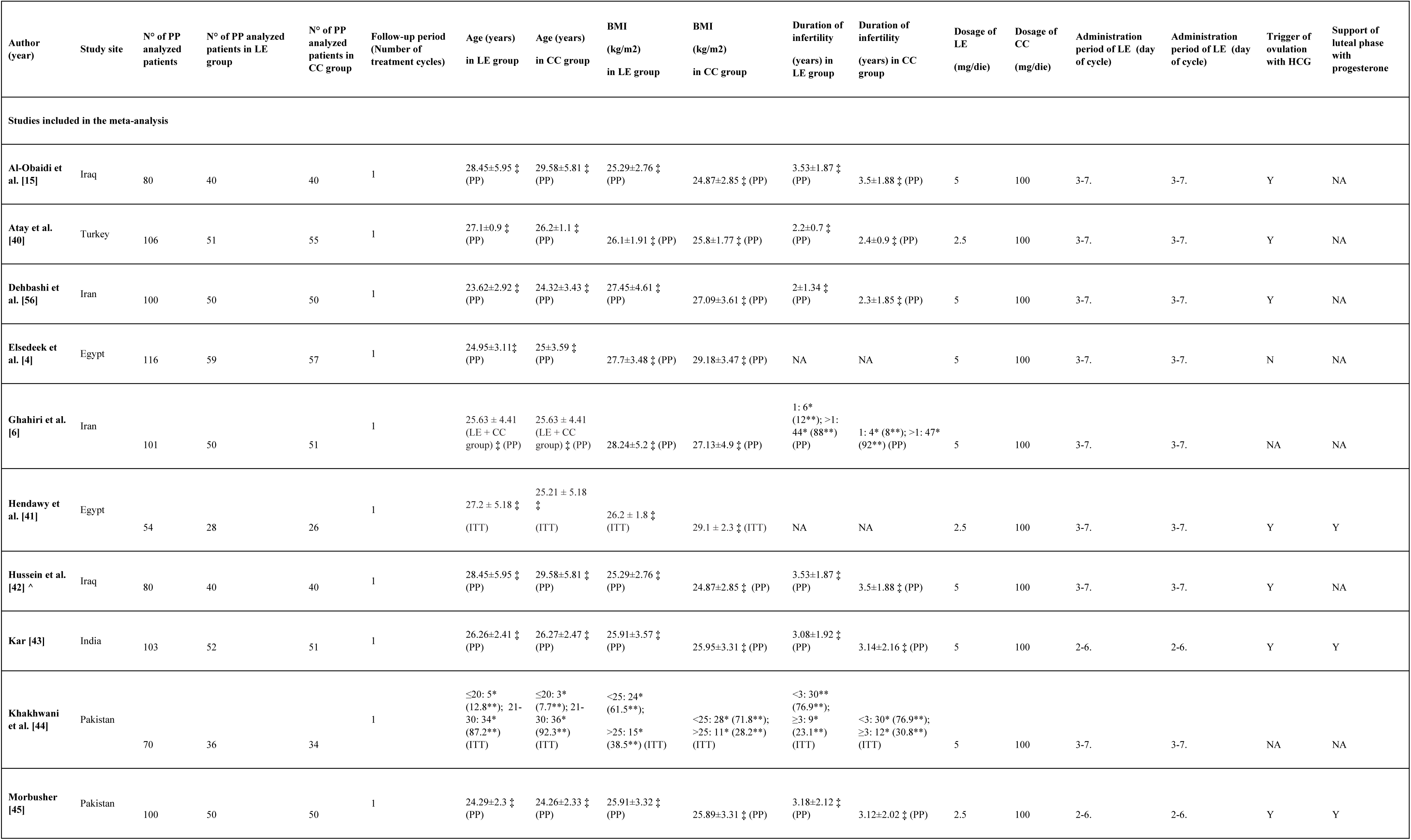
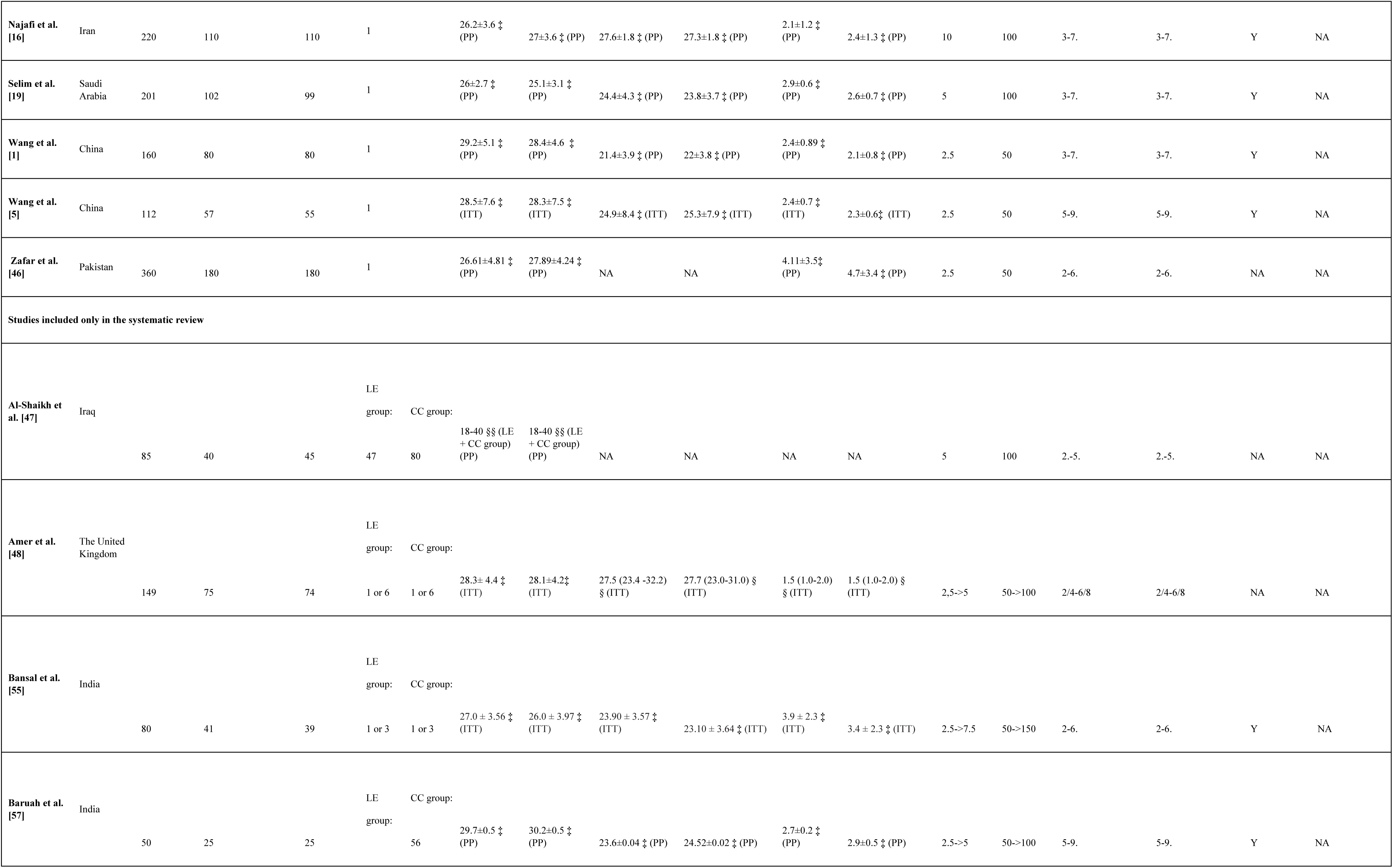
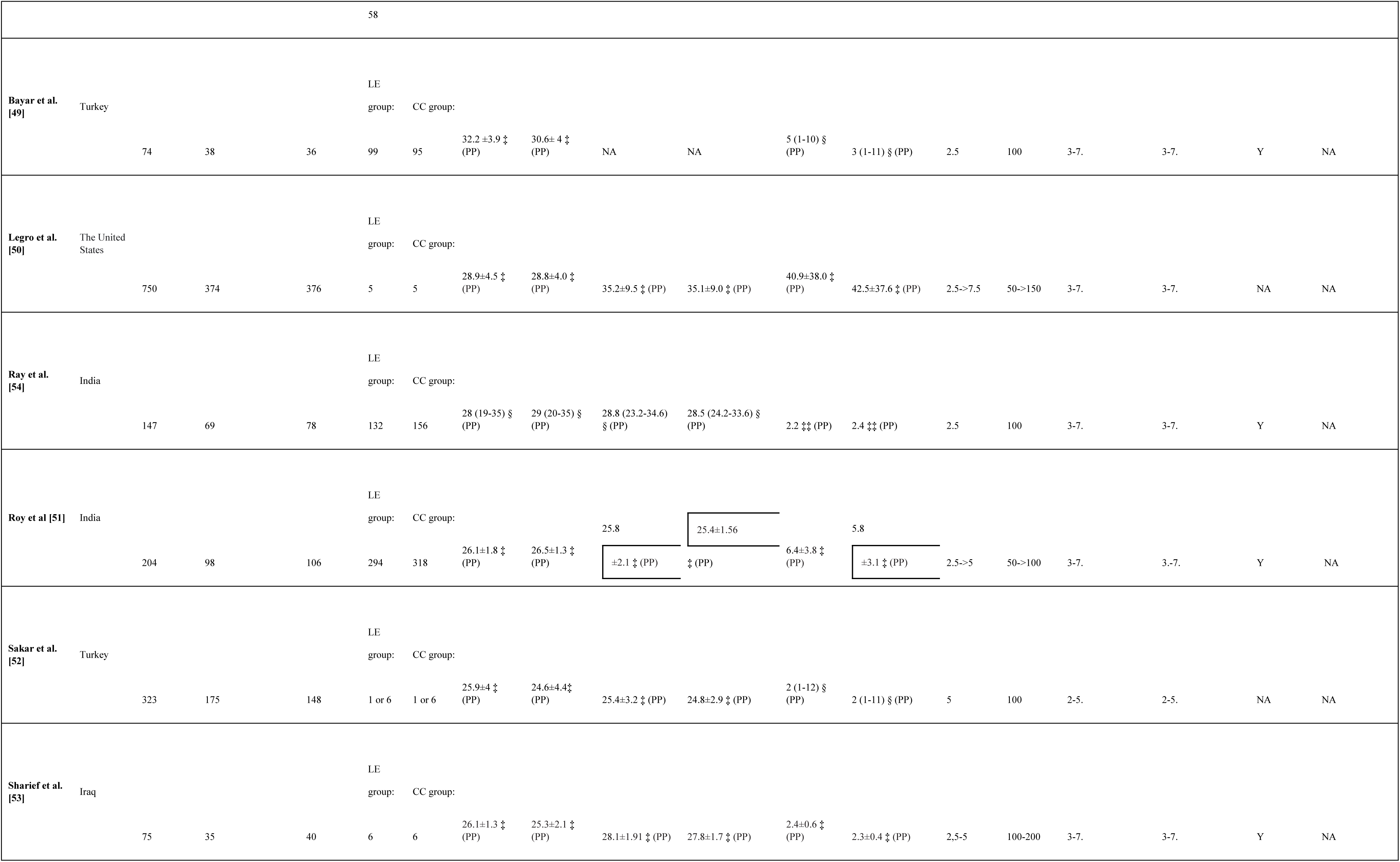
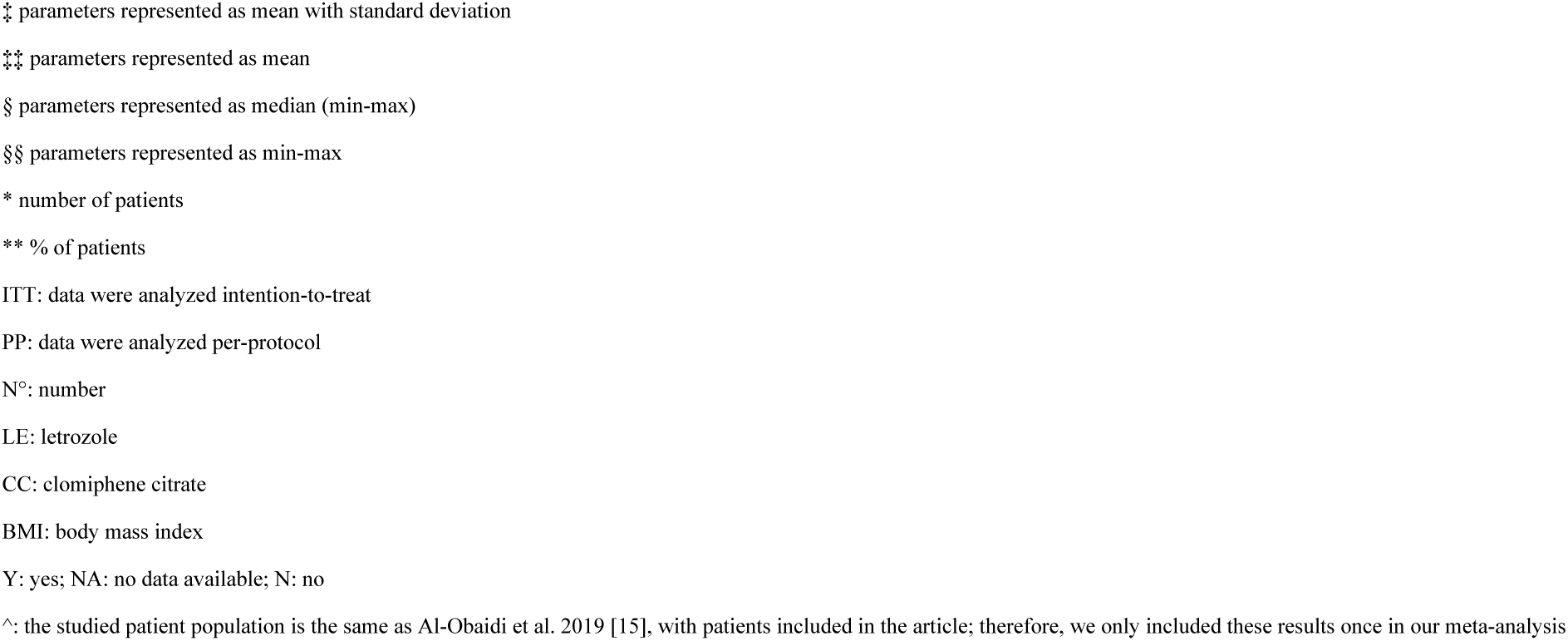
Basic characteristics of the studies included in the systematic review and meta-analysis.

### Results of the quantitative analysis

On the basis of the results of 11 studies including 1,651 patients, **ET** was significantly higher in LE group /Mean difference (MD)=1.70, CI: 0.55-2.86; I^2^=97%, p=0.008/ compared to CC group (**Fig 2A**). The subset analysis conducted on only ovulating patients showed thicker endometrium in LE group; however, this difference did not reach the level of statistical significance /MD=2.2, CI: -0.38-4.78; I^2^=97%, p=0.077/ (**S1 Fig**).

**Fig 2.**
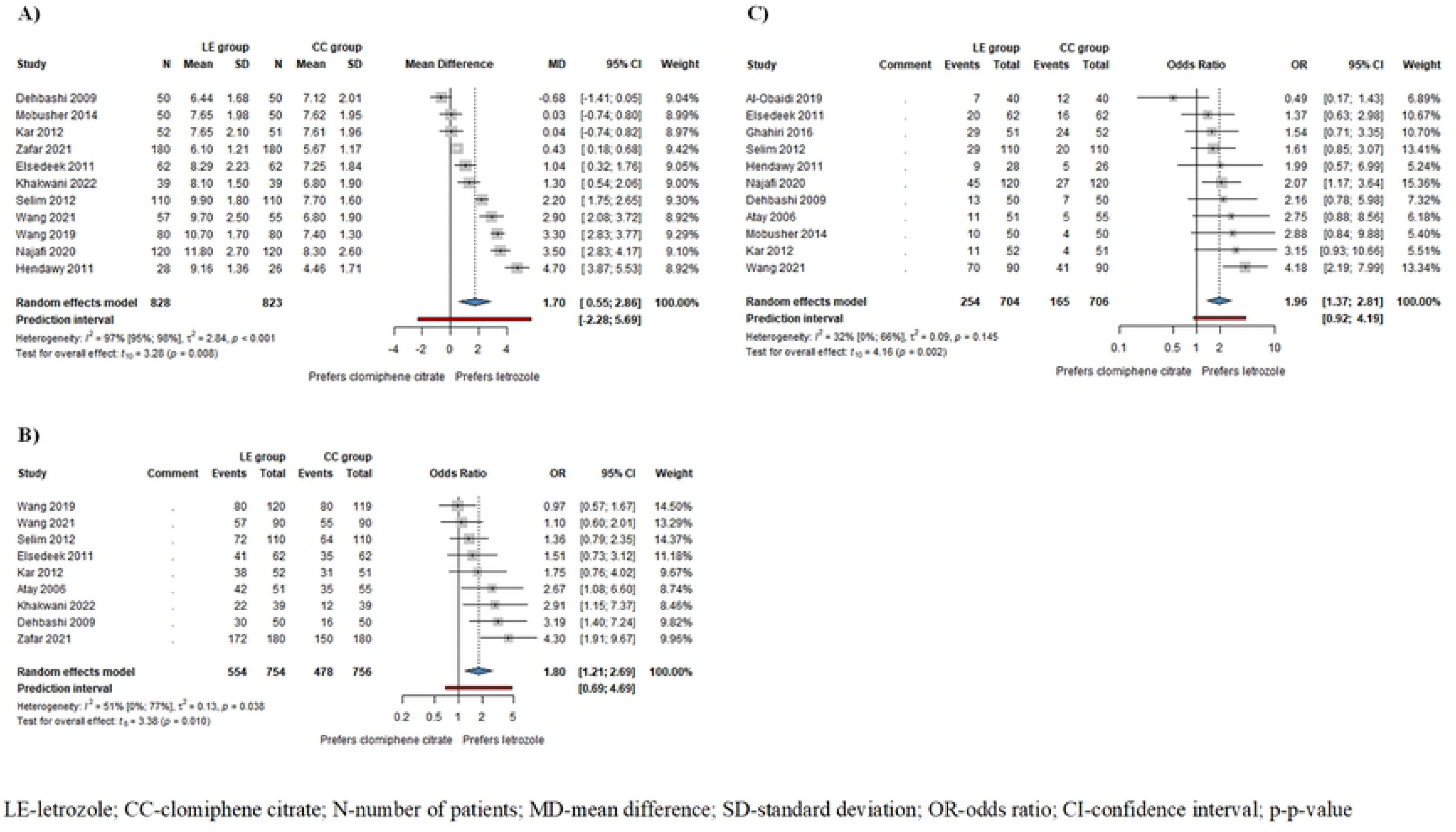
Forest plots for the following outcomes: A) endometrial thickness (ED - all patients: B) ovulation rate: O pregnancy rate - all patients

**Fig 2A) Forest plot for endometrial thickness (ET) - all patients.** Legend: LE-letrozole; CC-clomiphene citrate; N-number of patients; MD-mean difference; SD-standard deviation; CI-confidence interval; p-p-value.

**S1 Fig. Forest plot for endometrial thickness (ET) - ovulating patients only.** Legend: LE- letrozole; CC-clomiphene citrate; N-number of patients; MD-mean difference; SD-standard deviation; CI-confidence interval; p-p-value.

On the basis of the results of 9 studies including 1,510 patients, **odds of ovulation** and based on the results of 11 studies including 1410 patients, **odds of pregnancy** were significantly higher in LE patients /ovulation rate: odds ratio (OR)=1.80, CI: 1.21-2.69; I^2^=51%, p=0.010 (**Fig 2B**); pregnancy rate: OR=1.96, CI: 1.37-2.81; I^2^=32%, p=0.002 (**Fig 2C**)/ compared to CC patients. In patients who ovulated as a result of the drug therapy used, odds of pregnancy were also higher in LE group, but this difference did not reach the level of statistical significance /OR=1.65, CI: 0.40-6.76; I^2^=56%, p=0.337/ (**Fig S5**).

**Fig 2B) Forest plot for ovulation rate.** Legend: LE-letrozole; CC-clomiphene citrate; N-number of patients; OR-odds ratio; CI-confidence interval; p-p-value.

**Fig 2C) Forest plot for pregnancy rate – all patients.** Legend: LE-letrozole; CC- clomiphene citrate; N-number of patients; OR-odds ratio; CI-confidence interval; p-p-value.

**S5 Fig. Forest plot for pregnancy rate - ovulating patients only.** Legend: LE-letrozole; CC-clomiphene citrate; N-number of patients; OR-odds ratio; CI-confidence interval; p-p-value.

On the basis of the results of 3 studies including 460 patients, **RI of subendometrial arteries** was significantly lower in LE patients /MD=-0.15, CI: -0.27- -0.04; I^2^=92%, p=0.030/ compared to CC patients (**Fig 3A**). **PI of subendometrial arteries** was also lower in the LE group /MD=-0.17, CI: -0.81-0.47; I^2^=95%, p=0.372/, but this difference did not reach the level of statistical significance (**Fig 3B**).

**Fig 3.**
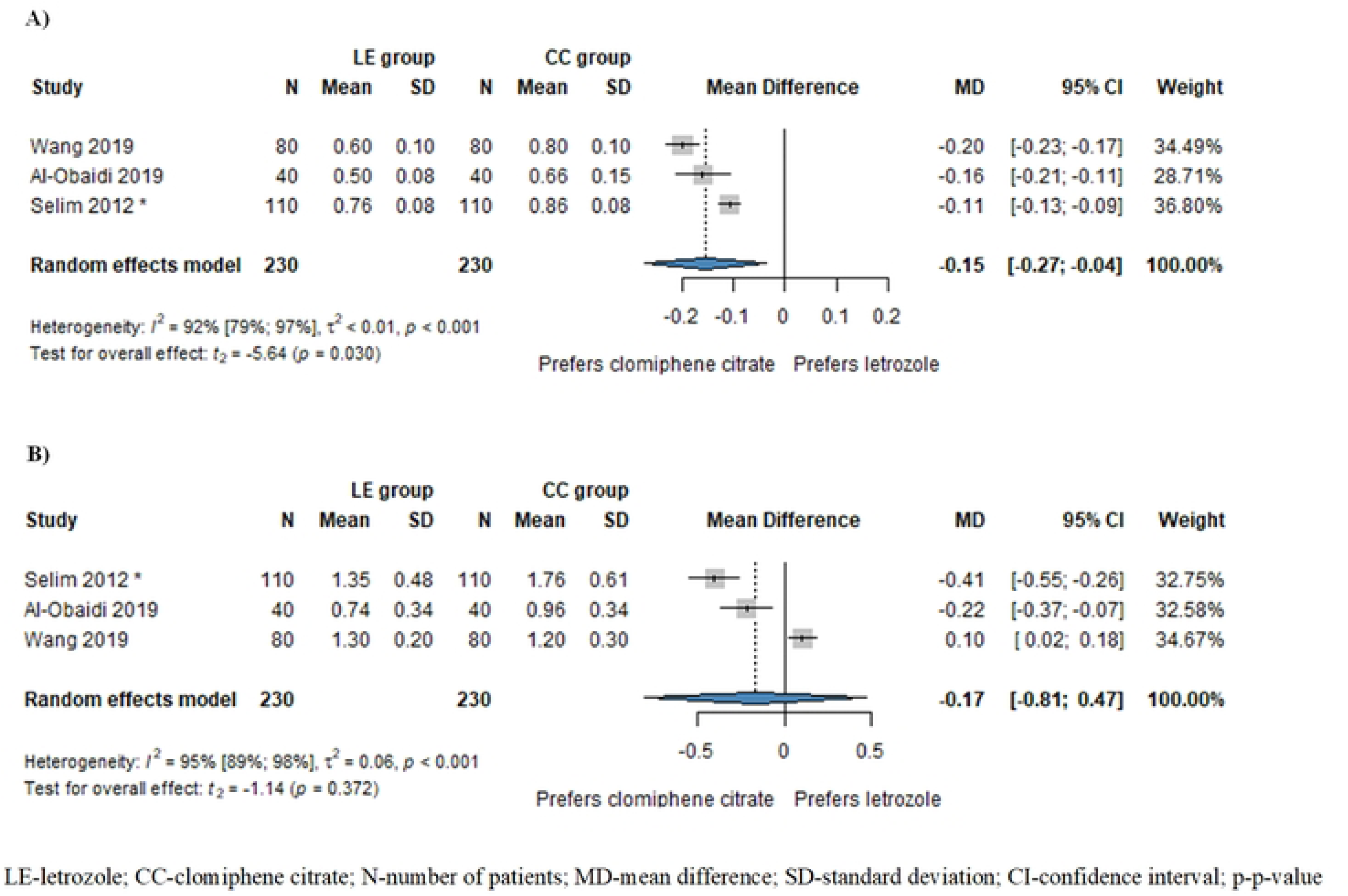
Forest plots for subendomendometrial circulation: A) resistance index (RI) of subendometrial arteries; B) pulsatility index (PI) of subendometrial arteries

**Fig 3A) Forest plot for subendomendometrial circulation - resistance index (RI) of subendometrial arteries.** Legend: LE-letrozole; CC-clomiphene citrate; N-number of patients; MD-mean difference; SD-standard deviation; CI-confidence interval; p-p-value.

**Fig 3B) Forest plot for subendomendometrial circulation - pulsatility index (PI) of subendometrial arteries.** Legend: LE-letrozole; CC-clomiphene citrate; N-number of patients; MD-mean difference; SD-standard deviation; CI-confidence interval; p-p-value.

On the basis of the results of 8 studies including 253 patients, **odds of multiple pregnancies** were higher in CC patients /multiple pregnancies: OR=0.41, CI: 0.12-1.35; I^2^=0%, p=0.119/ compared to LE patients **(S6 Fig)**, but this difference did not reach the level of statistical significance.

**S6 Fig. Forest plot for multiple pregnancy rate.** Legend: LE-letrozole; CC-clomiphene citrate; N-number of patients; OR-odds ratio; CI-confidence interval; p-p-value.

On the basis of the results of 4 studies including 160 patients, there was no difference in the **miscarriage** rate /OR=0.62, CI: 0.19-1.98; I^2^=0%, p=0.278/ (**S7 Fig**).

**S7 Fig. Forest plot for miscarriage rate.** Legend: LE-letrozole; CC-clomiphene citrate; N-number of patients; OR-odds ratio; CI-confidence interval; p-p-value.

There was no significant difference between groups in the **number** of **follicles** /MD=-0.40, CI: -0.84-0.03; I^2^=91%, p=0.066/ based on the results of 9 studies including 1264 patients (**S2 Fig**) and the **diameter** of **follicles** /MD=0.58, CI: -0.17-1.32; I^2^=45%, p=0.092/ based on the results of 4 studies including 660 patients (**S4 Fig**) although more but smaller follicles were seen in CC group. In the subset analysis of the number of follicles conducted in patients who only ovulated, there was also no significant difference between the two groups /MD=-0.80 CI: -2.48-0.89; I^2^= 80%; p=0.179/ **(S3 Fig**).

**S2 Fig. Forest plot for number of dominant follicles – all patients.** Legend: LE-letrozole; CC-clomiphene citrate; N-number of patients; MD-mean difference; SD-standard deviation. **CI-confidence interval; p-p-value.** Legend: LE-letrozole; CC-clomiphene citrate; N-number of patients; MD-mean difference; SD-standard deviation.

**S4 Fig. Forest plot for diameter of dominant follicles.** Legend: LE-letrozole; CC- clomiphene citrate; N-number of patients; MD-mean difference; SD-standard deviation.

**S3 Fig. Forest plot for number of dominant follicles - ovulating patients only.** Legend: LE-letrozole; CC-clomiphene citrate; N-number of patients; MD-mean difference; SD-standard deviation.

On the basis of the results of 3 studies including 563 patients, the **odds** were higher **for monofollicular development** and lower **for multifollicular development** in LE patients/monofollicular development: OR=1.99, CI: 0.62-6.34; I^2^=51%, p=0.126 (**S8 Fig**); in the case of multifollicular development: OR=0.50, CI: 0.16-1.61; I^2^=51%, p=0.126 (**S9 Fig**)/ compared to CC patients, but this difference did not reach the level of statistical significance.

**S8 Fig. Forest plot for monofollicular development rate.** Legend: LE-letrozole; CC- clomiphene citrate; N-number of patients; OR-odds ratio; CI-confidence interval; p-p-value.

**S9 Fig. Forest plot for multifollicular development rate**. Legend: LE-letrozole; CC- clomiphene citrate; N-number of patients; OR-odds ratio; CI-confidence interval; p-p-value.

### Results of the qualitative analysis

Results and main conclusions of the studies only comprised in the systematic review are detailed in **Table 2**.

**Table 2.**
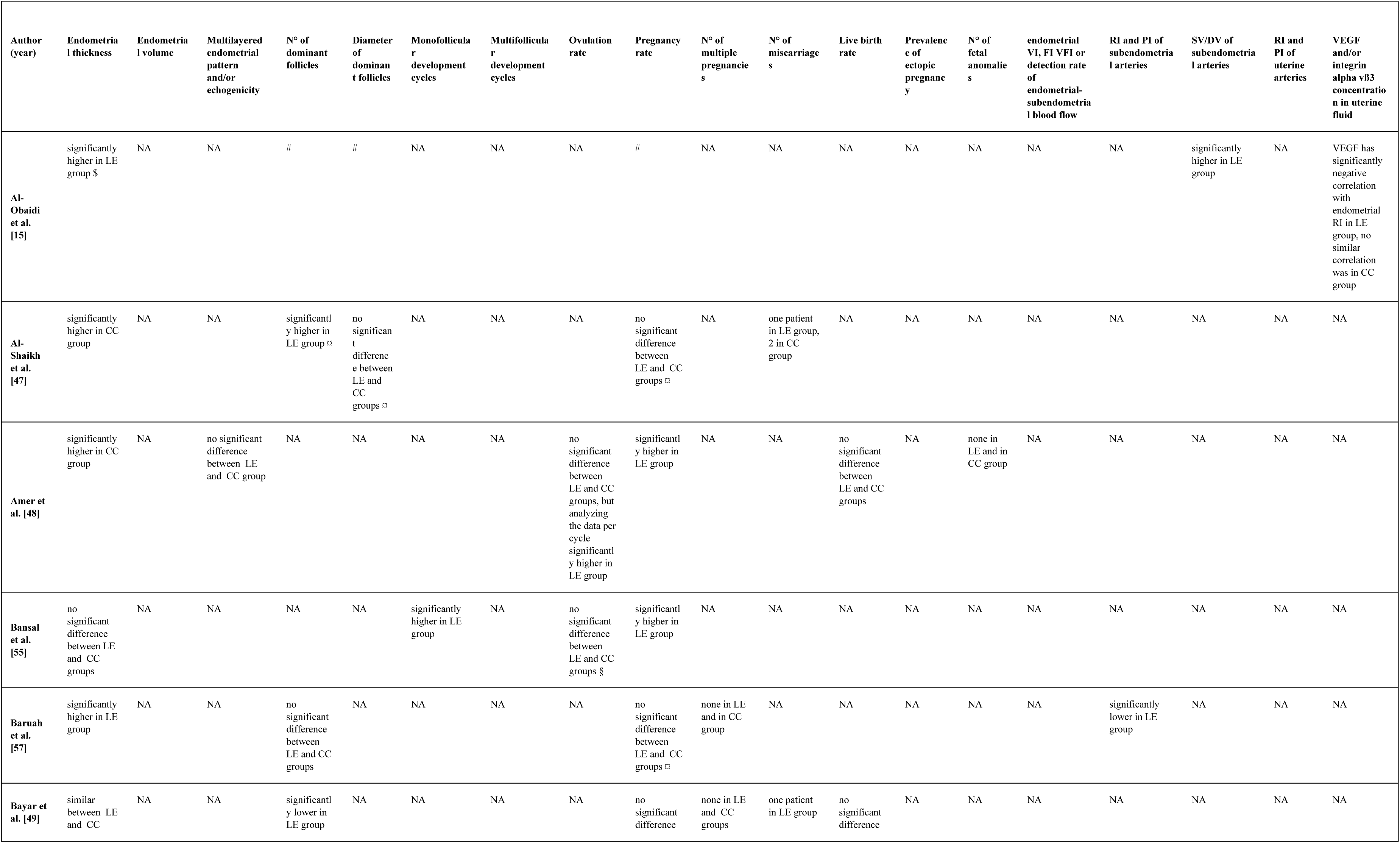
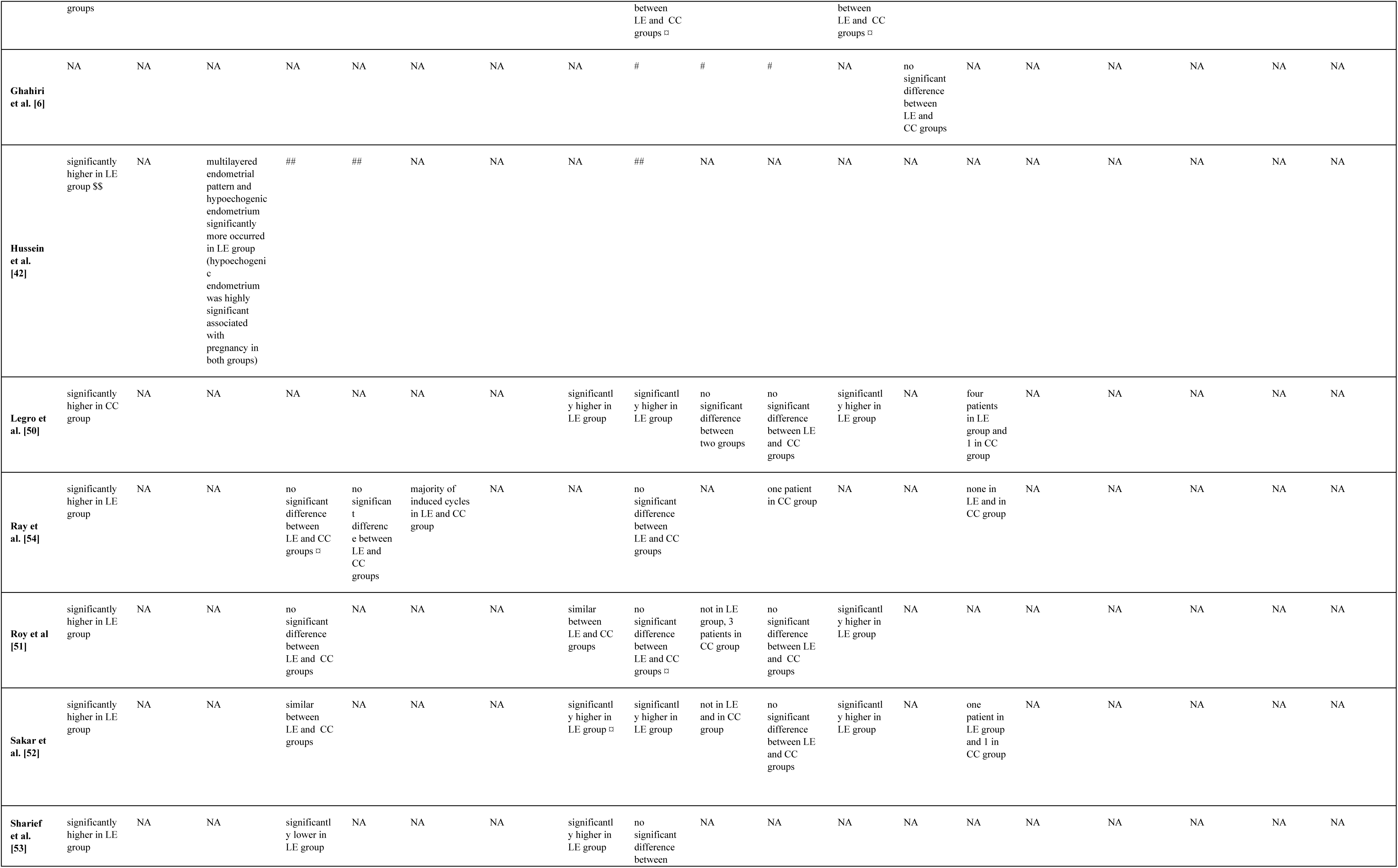
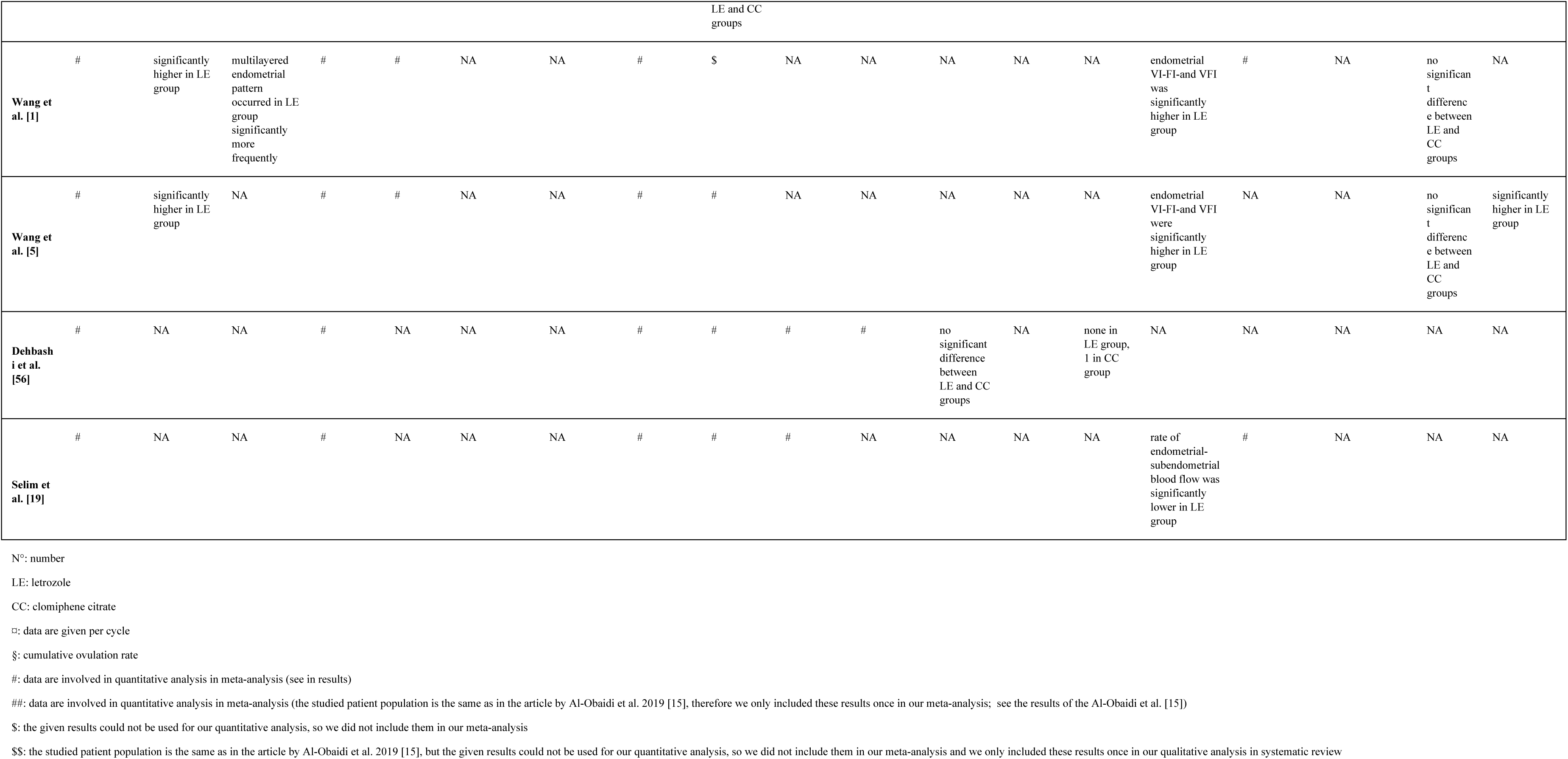
Main conclusions of the studies only included in the systematic review.

### Quality assessment

The results of the quality assessment are listed in the Summary of Findings Table (**S1 Table** in the Supplement).

### Publication bias

No evidence of publication bias was detected (**S10-14 Figs** in the Supplement).

The funnel plots do not indicate publication bias and the result of Egger’s test was not significant, which means publication bias could not be detected. Significant heterogeneity was observed for ET, number of dominant follicles and diameter of dominant follicles, RI of subendometrial artery, and PI of subendometrial artery. Other outcome measures showed low or moderate heterogeneity.

### Risk of bias assessment

The results of the risk of bias assessment are shown **S15-70 Figs** in the Supplement.

## Discussion

PCOS and infertility caused by PCOS are huge problems worldwide, the treatment of which has not been solved. Intensive research is underway to find the right drug for ovulation induction; however, there is no comprehensive summary of the results of the studies so far. In our study, we summarized the results of the available publications and identified several positive properties of LE as an ovulation induction drug compared to the currently widely used CC.

Thicker endometrium is associated with better embryo implantation [20]. ET in our meta-analysis was significantly thicker in LE group compared to CC group. When we compared our result with previous meta-analyses by He et al. found that the data of the articles included in the analysis could not be pooled due to significant heterogeneity [58]. However, Gadalla et al. stated that the endometrium was significantly thinner in CC group [11]. Akinoso-Imran et al. found similar results after letrozole treatment [59]. Of the RCTs included in our qualitative analysis (**Table 2**), six found significantly thicker endometrium in LE group [15, 42, 51–54, 57], three found significantly thicker endometrium in CC group [47, 48, 50], and two found no significant difference between the two groups [49, 55]. In other studies, the endometrial volume was significantly higher [1, 5] significantly more multilayered endometrial pattern [1, 42] and hypoechogenic endometrium occurred [42] in LE group. In contrast, one found no significant difference between the two groups in terms of endometrial pattern and echogenicity [48].

Patients taking CC appear to have more but smaller follicles, which may reduce the chance of appropriate maturation, follicle rupture and increase the risk of hyperstimulation and the odds of multiple pregnancies [60]. We found that the number of dominant follicles was higher in patients taking CC, although the definition of dominant follicle differed in the articles (from 12 mm< to 18 mm =/<). The diameters of follicles did not differ significantly; however, the follicles in LE group were larger. When we compared our results with previous meta-analyses, we found that He et al. observed significantly fewer mature follicles per cycle in patients taking letrozole [58]. In another study, the standardized mean difference revealed no statistically significant difference between patients treated with LE or CC, but there were also fewer mature follicles in letrozole group [59]. Of the RCTs included in our qualitative analysis (**Table 2**), one found significantly higher numbers of dominant follicles in LE group [47], two found significantly lower numbers in LE group [48, 49], and four found no significant difference between the two groups [51, 52, 54, 57]. As for the diameter of the dominant follicles, two found no significant difference between the two groups [47, 54].

Ovulation is essential for fertilization. We have previously presented that patients in LE group had significantly higher odds of ovulation than in CC group. In comparison of our result with previous meta-analyses, He et al. [58], Tsiami et al. [61] and Zhuo et al. [62] reported the same result. Wang et al. pointed out that the ovulation rate was significantly better when patients were taking LE instead of CC [63]. In contrast, Roque et al. [64] and Gadalla et al. [11] reported that the odds of ovulation were similar in letrozole group and in CC group.

Akinoso-Imran et al. also found that letrozole treatment caused a significantly higher ovulation rate compared to CC [59]. Shifu Hu et al. had similar results [65]. Of the RCTs included in our qualitative analysis (**Table 2**), three found significantly higher ovulation rates in LE group [50, 52, 53], two found no significant difference between the two groups [51, 55]. One found no significant difference in ovulation rate, but when analyzing the data per cycle found, it reported significantly higher in LE group [48]. Another one found the monofollicular development significantly higher [55] in LE group and one found the same majority of cycles included in both groups [54].

It would be good if more women with PCOS increased their chances of getting pregnant. We found that the odds of pregnancy were significantly higher in patients treated with letrozole than in CC group. In comparison of our result with previous meta-analyses, four were found to have significantly higher pregnancy rates [59, 62–64] and three were reported to have significantly higher odds of live birth [62–64] after letrozole treatment. Gadalla et al. [11] and Hu et al. [65] described similar differences between the two groups. In contrast, He et al. found no significant difference in the odds of pregnancy between the two groups [58]. Of the RCTs included in our qualitative analysis (**Table 2**), four found significantly higher pregnancy rates in LE group [48, 50, 52, 55], but the other six found no significant difference between the two groups [47, 49, 51, 53, 54, 57]. Three found significantly higher live birth rates in LE group [50–52], and three found no significant difference between the two groups [48, 49, 56].

Multiple pregnancy increases the odds of almost every potential complication of singleton pregnancy (e.g. preterm birth, fetal growth restriction, etc.) [66]. In our research, the odds of single pregnancies were higher amongst letrozole patients, and more multiple pregnancies occurred amongst CC patients, although this difference did not reach the level of statistical significance. Four previous meta-analyses came to a similar non-significant conclusion [58, 59, 63, 65]. In three of the RCTs included in our qualitative analysis (**Table 2**), no multiple pregnancies occurred in either group [49, 52, 57]. One found no multiple pregnancy in LE group, and three in CC group [51]. One found no significant difference between the two groups [50].

The aim should not only be to achieve pregnancy, but also to carry as many pregnancies to term as possible among women with PCOS. In our study, the odds of miscarriage were higher in CC group, but this difference did not reach the level of statistical significance. Our results are consistent with previous meta-analyses in the literature. [58, 59, 63–65]. Of the RCTs included in our qualitative analysis (**Table 2**), three found no significant difference in the number of miscarriages between the two groups [50–52], other studies found no difference or one or two in groups [47, 49, 54]. In the prevalence of ectopic pregnancy, one found no significant difference between the two groups [57]. Fetal anomalies were not found in either group in two RCTs [48, 54], Legro et al. found four in LE group, one in CC group [50], one found one in LE group, one in CC group [52], one other found no cases in LE group, one in CC group [56].

The RI of subendometrial arteries was significantly lower in LE group than in CC group. The PI of subendometrial arteries was also lower in patients treated with letrozole, but this result was not statistically significant. With lower RI and PI, intrauterine blood supply is better, and there may be a greater chance of improved intrauterine embryonic development. Wang et al. in their RCT found no difference in PI values between the LE or the CC group after Diane-35 and metformin pretreatment (therefore, we did not include these articles in our meta-analysis); however, the RI of subendometrial arteries was also significantly lower in LE group [67]. One RCT examined PI and RI of spiral arteries for six months; RI and PI were significantly lower in letrozole group [68]. Of the RCTs included in our qualitative analysis (**Table 2**), one also found significantly lower RI and PI of subendometrial arteries in LE group (they used an increasing drug dose and examined several cycles, so they were not included in our meta-analysis) [57]. Two found significantly higher endometrial vascularization index, flow index and vascularization flow index in LE group, and no significant difference in RI and PI of uterine arteries between the two groups [1, 5]. One found that the detection rate of endometrial-subendometrial blood flow was significantly lower in LE group [19]. One found significantly higher systolic velocity/diastolic velocity of subendometrial arteries and a significantly negative correlation in vascular endothelial growth factor concentration with endometrial RI in LE group, although there was no similar correlation in CC group [15]. One found significantly higher vascular endothelial growth factor and integrin alpha vß3 concentrations in uterine fluid in LE group [5].

We obtained results similar to the majority of previous meta-analyses, with the difference that we also highlighted that a better effect can be achieved with the use of LE compared to CC, even in one cycle. We are the first to use a meta-analysis to confirm the result that the use of LE might be beneficial for subendometrial ciculation compared to CC, which may explain the significant increase in the pregnancy rate among women with PCOS treated with letrozole. The previously mentioned changes in the subendometrial circulation are very early changes after one cycle, which may explain the differences in significance between RI and PI values. With longer-term use, the differences might also become significant for the PI values.

### Strengths and limitations

We used only randomized trials for our meta-analysis. The unique quality of this meta-analysis is that we separated and systematized the results of previous studies on women with PCOS treated with LE or CC according to whether the patients were followed for one or more cycles and whether they received a fixed or increasing drug dose used to induce ovulation. In this way, in contrast to previous meta-analyses, we have reduced the possibility of distortion that could be resulted by a quantitative analysis after combining the above research results.

For a quantitative analysis, we found sufficient data only from research studies that used fix doses of the drugs over one cycle.

Only a moderate number of studies could be included because the results of patients examined over one cycle could not be combined with the total results of patients examined over several cycles, nor with the results of patients whose medication dose was increased during the examination, if necessary, in order to provide accurate data. Furthermore, not all eligible studies used the same drug dose of LE or CC.

Another limitation is the presence of moderate and high risk of bias in some of the domains.

### Implications for practice and research

The significance of immediate application of research findings in clinical practice cannot be underestimated [69]. On the basis of the results of research findings we have analyzed, we can claim that LE should be considered for introduction into clinical practice worldwide among women with PCOS as a first-line oral ovulation induction drug. On the basis of our very encouraging results, we suggest further prospective data collection on subendometrial arterial circulation in women with PCOS during their treatment with letrozole for OI.

In the future, in order to provide more accurate data, it would be important to conduct as many research studies with the same number of cycles as possible and to accurately record the effects observed with each drug dose, to the extent that the dose of LE and CC is increased during the study.

## Conclusion

Our results show that letrozole is an effective therapeutic option for the treatment OI in patients with PCOS. Comparing LE with CC, which is the gold standard drug for OI, we can claim that the administration of LE resulted higher ovulation rates, thicker endometrium, lower RI of subendometrial arteries, and higher pregnancy rates. Lower RI in the subendometrial arteries can not only increase the chance of embryo implantation but also improve embryo development. For the above reasons, we recommend that LE should be considered as a first-line drug for ovulation induction.

## Data Availability

All relevant data are within the manuscript and its Supporting Information files.

## Acknowledgments

None.

## CRediT author contribution statement

**RZSV:** conceptualisation, project administration, methodology, formal analysis, writing – original draft; **AMG:** conceptualisation, data curation, writing - review & editing; **FAM:** conceptualisation; supervision; writing – original draft; **NÁ:** conceptualisation, writing – review & editing; **PH:** conceptualisation, writing - review & editing; **EZF:** conceptualisation, data curation, writing - review & editing; **PF:** conceptualization and design of the study, data curation, writing - review & editing; **SZK-D:** conceptualisation, data curation, writing - review & editing; **SZV**: conceptualisation, formal analysis, visualization, writing – review & editing; **JRH:** conceptualisation, writing -review & editing; **LS:** conceptualisation; supervision; writing – original draft

## Ethical approval

No ethical approval was required for this systematic review with meta-analysis, as all data were already published in peer-reviewed journals. No patients were involved in the design, conduct, or interpretation of our study.

The datasets used in this study can be found in the full-text articles included in the systematic review and meta-analysis.

## Supporting Information

**S1 Table. Summary of Findings Table.** Legend: CI: confidence interval, MD: mean difference, OR: odds ratio

**S2 Table. PRISMA 2020 Checklist.** Legend: **-**

**S2 Fig. Forest plot for number of dominant follicles – all patients.** Legend: LE-letrozole; CC-clomiphene citrate; N-number of patients; MD-mean difference; SD-standard deviation; CI-confidence interval; p-p-value.

**S3 Fig. Forest plot for number of dominant follicles – ovulating patients only.** Legend: LE-letrozole; CC-clomiphene citrate; N-number of patients; MD-mean difference; SD-standard deviation; CI-confidence interval; p-p-value.

**S4 Fig. Forest plot for diameter of dominant follicles.** Legend: LE-letrozole; CC- clomiphene citrate; N-number of patients; MD-mean difference; SD-standard deviation; CI- confidence interval; p-p-value.

**S8 Fig: Forest plot for monofollicular development rate.** Legend: LE-letrozole; CC- clomiphene citrate; N-number of patients; OR-odds ratio; CI-confidence interval; p-p-value.

**S9 Fig: Forest plot for multifollicular development rate.** Legend: LE-letrozole; CC- clomiphene citrate; N-number of patients; OR-odds ratio; CI-confidence interval; p-p-value.

**S10 Fig. Funnel plot for endometrial thickness (ET) – all patients.** Legend: -

**S11 Fig. Funnel plot for ovulation rate.** Legend: -

**S12 Fig. Funnel plot for pregnancy rate – all patients.** Legend: -

**S13 Fig. Funnel plot for resistance index (RI) of subendometrial arteries.** Legend: -

**S14 Fig. Funnel plot for pulsatility index (PI) of subendometrial arteries.** Legend: -

**S15 Fig. Risk of bias assessment of the studies included in the meta-analysis assessing endometrial thickness (ET)** [1, 4, 5, 16, 19, 41, 43–46, 56] **using the revised tool for assessing risk of bias in randomized trials (Rob 2).** Legend: +: low risk; !: some concerns; -: high risk

**S16 Fig. Risk of bias assessment of the studies included in the meta-analysis assessing endometrial thickness (ET)** [1, 4, 5, 16, 19, 41, 43–46, 56] **broken down to tools, shown in percentage.**

**S17 Fig. Risk of bias assessment of the studies included in the meta-analysis assessing number of dominant follicles** [1, 4, 5, 15, 16, 19, 40, 41, 56] **using the revised tool for assessing risk of bias in randomized trials (Rob 2).** Legend: +: low risk; !: some concerns; - : high risk

**S18 Fig. Risk of bias assessment of the studies included in the meta-analysis assessing number of dominant follicles** [1, 4, 5, 15, 16, 19, 40, 41, 56] **broken down to tools, shown in percentage.**

**S19 Fig. Risk of bias assessment of the studies included in the meta-analysis assessing diameter of dominant follicles** [1, 5, 15, 16] **using the revised tool for assessing risk of bias in randomized trials (Rob 2).** Legend: +: low risk; !: some concerns; -: high risk

**S20 Fig. Risk of bias assessment of the studies included in the meta-analysis assessing diameter of dominant follicles** [1, 5, 15, 16] **broken down to tools, shown in percentage.**

**S21 Fig. Risk of bias assessment of the studies included in the meta-analysis assessing mono-and multifollicular development** [43, 45, 46] **rate using the revised tool for assessing risk of bias in randomized trials (Rob 2).** Legend: +: low risk; !: some concerns; - : high risk

**S22 Fig. Risk of bias assessment of the studies included in the meta-analysis assessing mono-and multifollicular development** [43, 45, 46] **rate broken down to tools, shown in percentage.**

**S23 Fig. Risk of bias assessment of the studies included in the meta-analysis assessing ovulation rate** [1, 4, 5, 19, 42–44, 46, 56] **using the revised tool for assessing risk of bias in randomized trials (Rob 2).** Legend: +: low risk; !: some concerns; -: high risk

**S24 Fig. Risk of bias assessment of the studies included in the meta-analysis assessing ovulation rate** [1, 4, 5, 19, 42–44, 46, 56] **broken down to tools, shown in percentage.**

**S25 Fig. Risk of bias assessment of the studies included in the meta-analysis assessing pregnancy rate** [4–6, 15, 16, 19, 40, 42, 43, 45, 56] **using the revised tool for assessing risk of bias in randomized trials (Rob 2).** Legend: +: low risk; !: some concerns; -: high risk

**S26 Fig. Risk of bias assessment of the studies included in the meta-analysis assessing pregnancy rate** [4–6, 15, 16, 19, 40, 42, 43, 45, 56] **broken down to tools, shown in percentage.**

**S27 Fig. Risk of bias assessment of the studies included in the meta-analysis assessing single and multiple pregnancy rate** [6, 16, 19, 40, 41, 45, 56] **using the revised tool for assessing risk of bias in randomized trials (Rob 2).** Legend: +: low risk; !: some concerns; - : high risk

**S28 Fig. Risk of bias assessment of the studies included in the meta-analysis assessing single and multiple pregnancy rate** [6, 16, 19, 40, 41, 45, 56] **broken down to tools, shown in percentage.**

**S29 Fig. Risk of bias assessment of the studies included in the meta-analysis assessing rate of miscarriage** [6, 16, 43, 56] **using the revised tool for assessing risk of bias in randomized trials (Rob 2).** Legend: +: low risk; !: some concerns; -: high risk

**S30 Fig. Risk of bias assessment of the studies included in the meta-analysis assessing rate of miscarriage** [6, 16, 43, 56] **broken down to tools, shown in percentage.**

**S31 Fig. Risk of bias assessment of the studies included in the meta-analysis assessing resistance index (RI) of subendometrial arteries** [1, 15, 19] **using the revised tool for assessing risk of bias in randomized trials (Rob 2).** Legend: +: low risk; !: some concerns; -: high risk

**S32 Fig. Risk of bias assessment of the studies included in the meta-analysis assessing resistance index (RI) of subendometrial arteries** [1, 15, 19] **broken down to tools, shown in percentage.**

**S33 Fig. Risk of bias assessment of the studies included in the meta-analysis assessing pulsatility index (PI) of subendometrial arteries** [1, 15, 19] **using the revised tool for assessing risk of bias in randomized trials (Rob 2).** Legend: +: low risk; !: some concerns; -: high risk

**S34 Fig. Risk of bias assessment of the studies included in the meta-analysis assessing pulsatility index (PI) of subendometrial arteries** [1, 15, 19] **broken down to tools, shown in percentage.**

**S35 Fig. Risk of bias assessment of the studies included in the systematic review assessing rate of endometrial thickness (ET)** [15, 42, 47–49, 51–55, 57] **using the revised tool for assessing risk of bias in randomized trials (Rob 2).** Legend: +: low risk; !: some concerns; -: high risk

**S36 Fig. Risk of bias assessment of the studies included in the systematic review assessing rate of endometrial thickness (ET)** [15, 42, 47–49, 51–55, 57] **broken down to tools, shown in percentage.**

**S37 Fig. Risk of bias assessment of the studies included in the systematic review assessing endometrial volume (EV)** [1, 5] **using the revised tool for assessing risk of bias in randomized trials (Rob 2).** Legend: +: low risk; !: some concerns; -: high risk

**S38 Fig. Risk of bias assessment of the studies included in the systematic review assessing endometrial volume (EV)** [1, 5] **broken down to tools, shown in percentage.**

**S39 Fig. Risk of bias assessment of the studies included in the systematic review assessing endometrial pattern and/or echogenicity** [1, 42, 48] **using the revised tool for assessing risk of bias in randomized trials (Rob 2).** Legend: +: low risk; !: some concerns; -: high risk

**S40 Fig. Risk of bias assessment of the studies included in the systematic review assessing endometrial pattern and/or echogenicity** [1, 42, 48] **broken down to tools, shown in percentage.**

**S41 Fig. Risk of bias assessment of the studies included in the systematic review assessing rate of number of dominant follicles** [47, 49, 51–54, 57] **using the revised tool for assessing risk of bias in randomized trials (Rob 2).** Legend: +: low risk; !: some concerns; -: high risk

**S42 Fig. Risk of bias assessment of the studies included in the systematic review assessing rate of number of dominant follicles** [47, 49, 51–54, 57] **broken down to tools, shown in percentage.**

**S43 Fig. Risk of bias assessment of the studies included in the systematic review assessing rate of diameter of dominant follicles** [47, 54] **using the revised tool for assessing risk of bias in randomized trials (Rob 2).** Legend: +: low risk; !: some concerns; - : high risk

**S44 Fig. Risk of bias assessment of the studies included in the systematic review assessing rate of diameter of dominant follicles** [47, 54] **broken down to tools, shown in percentage.**

**S45 Fig. Risk of bias assessment of the studies included in the systematic review assessing monofollicular development cycles** [54, 55] **using the revised tool for assessing risk of bias in randomized trials (Rob 2).** Legend: +: low risk; !: some concerns; -: high risk

**S46 Fig. Risk of bias assessment of the studies included in the systematic review assessing monofollicular development cycles** [54, 55] **broken down to tools, shown in percentage.**

**S47 Fig. Risk of bias assessment of the studies included in the systematic review assessing ovulation rate** [48, 50–53, 55] **using the revised tool for assessing risk of bias in randomized trials (Rob 2).** Legend: +: low risk; !: some concerns; -: high risk

**S48 Fig. Risk of bias assessment of the studies included in the systematic review assessing ovulation rate** [48, 50–53, 55] **broken down to tools, shown in percentage.**

**S49 Fig. Risk of bias assessment of the studies included in the systematic review assessing pregnancy rate** [47–55, 57] **using the revised tool for assessing risk of bias in randomized trials (Rob 2).** Legend: +: low risk; !: some concerns; -: high risk

**S50 Fig. Risk of bias assessment of the studies included in the systematic review assessing pregnancy rate** [47–55, 57] **broken down to tools, shown in percentage.**

**S51 Fig. Risk of bias assessment of the studies included in the systematic review assessing number of multiple pregnancies** [49–52, 57] **using the revised tool for assessing risk of bias in randomized trials (Rob 2).** Legend: +: low risk; !: some concerns; -: high risk

**S52 Fig. Risk of bias assessment of the studies included in the systematic review assessing number of multiple pregnancies** [49–52, 57] **broken down to tools, shown in percentage.**

**S53 Fig. Risk of bias assessment of the studies included in the systematic review assessing number of miscarriages** [47, 49–52, 54] **using the revised tool for assessing risk of bias in randomized trials (Rob 2).** Legend: +: low risk; !: some concerns; -: high risk

**S54 Fig. Risk of bias assessment of the studies included in the systematic review assessing number of miscarriages** [47, 49–52, 54] **broken down to tools, shown in percentage.**

**S55 Fig. Risk of bias assessment of the studies included in the systematic review assessing live birth rate** [48–52, 56] **using the revised tool for assessing risk of bias in randomized trials (Rob 2).** Legend: +: low risk; !: some concerns; -: high risk

**S56 Fig. Risk of bias assessment of the studies included in the systematic review assessing live birth rate** [48–52, 56] **broken down to tools, shown in percentage.**

**S57 Fig. Risk of bias assessment of the studies included in the systematic review assessing number of ectopic pregnancies [6] using the revised tool for assessing risk of bias in randomized trials (Rob 2).** Legend: +: low risk; !: some concerns; -: high risk

**S58 Fig. Risk of bias assessment of the studies included in the systematic review assessing number of ectopic pregnancies** [6] **broken down to tools, shown in percentage.**

**S59 Fig. Risk of bias assessment of the studies included in the systematic review assessing number of fetal anomalies** [48, 50, 52, 54, 56] **using the revised tool for assessing risk of bias in randomized trials (Rob 2).** Legend: +: low risk; !: some concerns; - : high risk

**S60 Fig. Risk of bias assessment of the studies included in the systematic review assessing number of fetal anomalies** [48, 50, 52, 54, 56] **broken down to tools, shown in percentage.**

**S61 Fig. Risk of bias assessment of the studies included in the systematic review assessing endometrial vascularization index (VI), flow index (FI), vascularization flow index (VFI) and detection rate of endometrial-subendometrial blood flow** [1, 5, 19] **using the revised tool for assessing risk of bias in randomized trials (Rob 2).** Legend: +: low risk; !: some concerns; -: high risk

**S62 Fig. Risk of bias assessment of the studies included in the systematic review assessing endometrial vascularization index (VI), flow index (FI), vascularization flow index (VFI) detection rate of endometrial-subendometrial blood flow** [1, 5, 19] **broken down to tools, shown in percentage.**

**S63 Fig. Risk of bias assessment of the studies included in the systematic review assessing systolic velocity (SV)/diastolic velocity (DV) of subendometrial arteries** [15] **using the revised tool for assessing risk of bias in randomized trials (Rob 2).** Legend: +: low risk; !: some concerns; -: high risk

**S64 Fig. Risk of bias assessment of the studies included in the systematic review assessing systolic velocity (SV)/diastolic velocity (DV) of subendometrial arteries** [15]**broken down to tools, shown in percentage.**

**S65 Fig. Risk of bias assessment of the studies included in the systematic review assessing resistance index (RI) and pulsatility index (PI) of uterine arteries** [1, 5] **using the revised tool for assessing risk of bias in randomized trials (Rob 2).** Legend: +: low risk; !: some concerns; -: high risk

**S66 Fig. Risk of bias assessment of the studies included in the systematic review assessing resistance index (RI) and pulsatility index (PI) of uterine arteries** [1, 5] **broken down to tools, shown in percentage.**

**S67 Fig. Risk of bias assessment of the studies included in the systematic review assessing vascular endothelial growth factor (VEGF) and/or integrin alpha vß3** [5, 15] **using the revised tool for assessing risk of bias in randomized trials (Rob 2).** Legend: +: low risk; !: some concerns; -: high risk

**S68 Fig. Risk of bias assessment of the studies included in the systematic review assessing vascular endothelial growth factor (VEGF) and/or integrin alpha vß3** [5, 15] **broken down to tools, shown in percentage.**

**S69 Fig. Risk of bias assessment of the studies included in the systematic review assessing resistance index (RI) and pulsatility index (PI) of subendometrial arteries** [57] **using the revised tool for assessing risk of bias in randomized trials (Rob 2)** Legend: +: low risk; !: some concerns; -: high risk

**S70 Fig. Risk of bias assessment of the studies included in the systematic review assessing resistance index (RI) and pulsatility index (PI) of subendometrial arteries** [57] **broken down to tools, shown in percentage.** Legend: +: low risk; !: some concerns; -: high risk

